# Wastewater-Based Epidemiological Modeling for Continuous Surveillance of COVID-19 Outbreak

**DOI:** 10.1101/2021.10.19.21265221

**Authors:** Mehrdad Fazli, Samuel Sklar, Michael D. Porter, Brent A. French, Heman Shakeri

## Abstract

Using wastewater surveillance as a continuous pooled sampling technique has been in place in many countries since the early stages of the outbreak of COVID-19. Since the beginning of the outbreak, many research works have emerged, studying different aspects of viral SARS-CoV-2 DNA concentrations (viral load) in wastewater and its potential as an early warning method. However, one of the questions that has remained unanswered is the quantitative relation between viral load and clinical indicators such as daily cases, deaths, and hospitalizations. Few studies have tried to couple viral load data with an epidemiological model to relate the number of infections in the community to the viral burden. This paper proposes a stochastic wastewater-based SEIR model to showcase the importance of viral load in the early detection and prediction of an outbreak in a community. We built three models based on whether or not they use the case count and viral load data and compared their simulations and forecasting quality. Our results demonstrate that a simple SEIR model based on viral load data can reliably predict the number of infections in the future. Therefore, wastewater-based surveillance is a promising way of monitoring the spread of COVID-19 and can provide city officials with timely information about the circulation of COVID-19 in the community.

## 1. Introduction

The sudden outbreak of the COVID-19 pandemic has exposed critical vulnerabilities in the healthcare system. In particular, hospitals located in hotspots were overrun with COVID-19 patients, placing unprecedented demands on healthcare workers and depleting hospital resources. In contrast, other hospitals canceled outpatient appointments and emptied their wards in preparation for a surge of patients that never materialized. In the aftermath of this pandemic, it is now clear that the appropriate regional allocation of medical resources (including health professionals and specialized equipment) is critically important in the fight to save lives in future outbreaks. Nevertheless, the most critical weakness exposed by the COVID-19 epidemic is that our current methods for accurately predicting the location of regional outbreaks are inadequate. Thus improved techniques are needed to forewarn of surges in case loads.

Accurately characterizing the spread of an infectious disease and the response of different population segments throughout an epidemic is critical to implementing effective mitigation strategies. In addition to the infectivity of the agent, the risk of an outbreak (i.e., a large percentage of the population becoming infected) is a function of (i) the heterogeneity of contacts of the human population, (ii) the prevalence of individual immunity, and (iii) the successful implementation of public health strategies. These factors are challenging to monitor due to incomplete observations of spatial contact patterns and infeasibility to perform individual testing for a large fraction of the population, which effectively leaves the asymptomatic and latent cases out of the equation. Moreover, the majority of current approaches for predicting the spread of COVID-19 rely on lagged indicators like positive test ratio and hospitalizations [1]. Given a purely reactive approach to testing, the earliest detection possible comes well after the onset of symptoms, typically from test results compiled by regional healthcare systems.

Currently, data collection for COVID-19 is conducted by governmental institutions and universities. Of particular interest to the scope of this paper, the Virginia Department of Health (VDH) provides an updated count of all cases, divided into a variety of geographic regions, which is freely accessible [2]. Additionally, Johns Hopkins University maintains a dashboard of cases across the United States, divided by county [3]. Because the existing dataset is reactive, computational modeling provides health systems with this much-needed warning.

The limitations of modeling input data described above suggest a need for more data sources, preferably those that can forewarn infection surges to medical systems. One of the most promising data collection methods is a pooled sampling approach, where collective samples are taken from an entire population. This method aims to estimate the overall infection rate while using a fraction of the resources that it would take to sample the whole population individually.

Numerous studies have established wastewater surveillance for monitoring COVID-19 as an effective pooled sampling technique. Two notable studies at both Yale University and in the Ishikawa and Toyama prefectures in Japan took samples of sewage sludge and municipal wastewater, respectively, and performed PCR analysis of the samples to calculate viral load [4], [5].

COVID-19 infected cases usually excrete SARS-CoV-2 RNA copies in their feces and urine [6] over the course of their illness. It has been shown that pre-symptomatic, asymptomatic, and mildly symptomatic (PAMS) patients can also shed SARS-CoV-2 [7]. According to [8], numerous studies reported positive viral load samples before the first infected case was clinically diagnosed in the community. This is particularly crucial for monitoring the COVID-19 pandemic as a large proportion of the infected population are PAMS [9]. This implies massive under-reporting that further accelerates the circulation of the infection in communities. Jones et al. [7] estimate that by the time that PAMS patients test positive for COVID-19, they are nearly as infectious as hospitalized patients and have a comparable viral load. Therefore, monitoring viral load in wastewater can precede lagging indicators such as clinically confirmed case counts, hospitalization, and deaths.

One major gap in wastewater surveillance is relating viral load data to case counts. Most of the studies relied only on correlation-based analysis to confirm that viral load can follow the pattern of prevalence data without providing a model to back-calculate the number of active shedders [10], [11], [12], [13], [14]. Few studies have employed a wastewater-based epidemiological model to explain the COVID-19 cases with the viral load data. Xiao et al. [15] used a transfer function in the form of beta distribution with unknown parameters to map the viral load data onto the reported case data in the Boston area. While providing a better picture of the linkage between viral load and active shedders, this method is incapable of explaining shedding mechanism like shedding profile of patients.

One of the first studies that introduced wastewater-based epidemiological model to associate viral load data with reported COVID-19 cases was done by McMahan et al. [16]. They considered an SEIR compartmental model with a time-varying shedding rate. They also considered the degradation of virus RNAs in sewer systems prior to sampling and viral load measurement and related the viral load to active cases on a given day in a formula. A more comprehensive epidemiological model is proposed by Nourbakhsh et al. [17] with more compartments to include severe cases, mild cases, and recovered cases who still shed the virus. They accounted for variation in reproduction number and delay in case count reporting. They showed that models that use both viral load and confirmed cases have better predictive power to forecast the infection incidences in three Canadian cities.

In this paper, we take a similar approach as McMahan et al. [16] and consider a stochastic epidemiological model that supplements the conventional reported cases with pooled samples from wastewater surveillance for assessing the overall SARS-CoV-2 viral burden at the community level. Additionally, we allow for stochasticity, imported infections and non-homogeneity in the mixing of the population. Then, we investigate the effectiveness and robustness of our modeling, comparing both simulations and predictions with the data.

## 2. Method

We consider a stochastic wastewater-based epidemiological model with four compartments (hidden states) of susceptible (S), exposed (E), infectious (I), and recovered/removed (R) (Fig. 1a). Stochastic compartment models are a type of partially observed Markov processes (POMP) models with stochastic transition rates. These models have been commonly applied to epidemiological data to model the evolution of epidemics, extract epidemiological indicators from the observed data, and forecast how the outbreak will unfold in the future [18].

**Figure 1:**
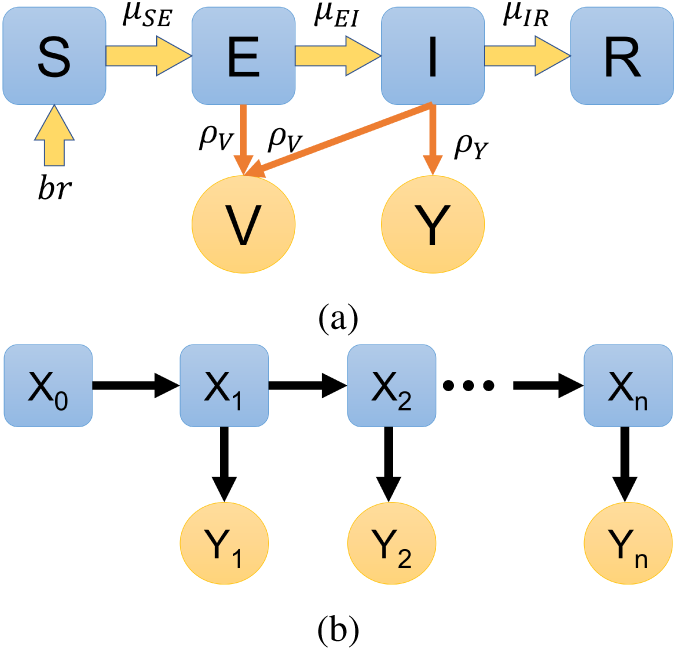
(a) Schematic of the SEIR model. (b) A simple Partially Observed Markov model

Partially Observed Markov Processes, also known as Hidden Markov Models (HMM), are one of the most powerful tools for time series analysis. They are defined over a sequence of latent (hidden) states and observed states. The simplest type of a POMP (HMM) model is the first-order HMM, which is shown in Fig. 1b. *X*_0:*N*_ are latent states, and *Y*_1:*N*_ are observed states, and 1 : *N* are the timesteps of the time series. This structure has two underlying assumptions that are crucial to our modeling:

- Given a specified model (known parameters), the latent state at time *t* depends only on the latent state at time *t* − 1.
- Given a specified model, the observed state at time *t* depends only on the hidden state at time *t*.

Now suppose that we have a sequence of *N* observations as *y*_1:*N*_ and a sequence of *N* + 1 hidden states as *x*_0:*N*_. Also, suppose that *θ* is the vector of parameters for our model. We can write the likelihood of the sequence of the observations considering our model as:

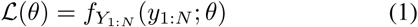

However, considering the aforementioned assumptions, the conditional probability rules, and the law of total probability, we can rewrite the likelihood as [19]:

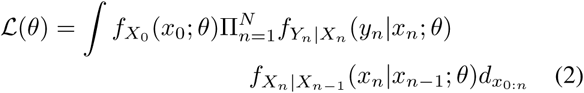

The right-hand side of Equation (2) is the initial density, one-step transition density, and measurement density in the order of appearance. To fully specify a POMP model, one has to specify a model for these three probability densities. Nevertheless, it is extremely difficult or impossible to define an analytical formula for the transition density in many problems. That is when the simulation-based inference is needed; Simulation-based inference attempts to simulate the transitions between the states instead of drawing from a known distribution. In this study, we use Iterated filtering [20], [21], to estimate the parameters of our model.

Our SEIR model has four latent states (*S, E, I, R*) and two observations (*V, Y*), namely viral load and reported cases per day. We define *N* := *S* + *E* + *I* + *R* as the total number of agents contributing to the dynamics of the epidemic at any given point in time. Moreover, let *br* be the birth rate and means transferring of agents from the inactive (non-contributing) population into the susceptible pool. We ignore death from the compartments due to the short epidemic period under study.

We assume agents in compartments *E* and *I* contribute to the viral load with a constant shedding rate, *ρ*_*V*_. Parameters *µ*_*SE*_, *µ*_*EI*_, and *µ*_*IR*_ are the rates at which agents transition from *S* to *E*, from *E* to *I* and from *I* to *R* respectively. These rates are inversely proportional to the expected time an agent spends at the respective compartment. We assume that *µ*_*EI*_ and *µ*_*IR*_ are constant over the duration of the study. The mean latent period that is reported by Centers for Disease Control and Prevention (CDC) in their summary reports on March 2021 [22] (time from exposure to symptom onset) is 6 days. Therefore, we set the *µ*_*EI*_ to 0.16 day^−1^.

The mean infectious period has been reported anywhere between 3.5 days to 11 days [23]. It is noteworthy that the mean infectious period greatly depends on the severity of the infection. However, it appears the average infectious period as a ballpark estimate is in the range of 6-8 days. Therefore, we set the *µ*_*IR*_ to be 0.13 day^−1^, i.e., the mean infectious period is 7.7 days. We also consider under-reporting in our model via parameter *ρ*_*Y*_. CDC in its report on July 2021 [24] suggests a reporting rate of 1*/*4.2 ≈ 0.24. However, as we focus on the beginning of the outbreak, the value of 0.14 seems more appropriate [9]. The parameter *µ*_*SE*_, defined in (15), represents the force of infection and measures the contact rate of infectious agents and susceptible agents. It will be a deciding factor on the diffusion of the infection through the community.

We start off by writing the equations for the number of agents at each compartment. Using equations 3-6, we update the state of the system:

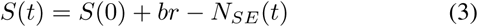

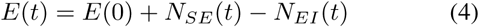

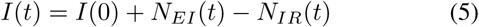

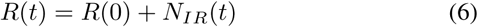

where *N*_*SE*_(*t*), *N*_*EI*_ (*t*), and *N*_*IR*_(*t*) are the number of agents transitioning from one compartment to the next one at a certain time. The initial state of the system is also defined via the following equations:

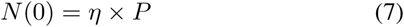

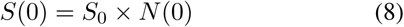

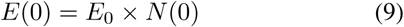

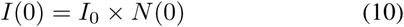

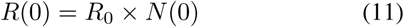

where *P* denotes the total population in the community under study and *η* is the fraction of them initially contributing to the spread and are present in one of the compartments at *t* = 0. *S*_0_, *E*_0_, *I*_0_, *R*_0_ are the initial value parameters specifying the fraction of agents at each compartment at the beginning of the study. Now we describe the dynamics of the transitions using the following ODEs:

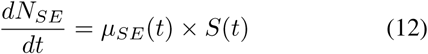

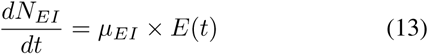

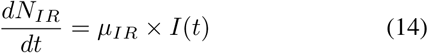

We allow for additional stochasticity into the force of infection *µ*_*SE*_ (following [25]) as,

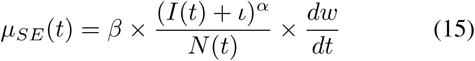

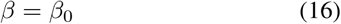

Where *β* = *β*_0_ is a constant transmission rate, *ι* is the imported infections from outside of the population, *α* is the mixing parameter with *α* = 1 representing a homogeneous mixing, and *w* is the Gamma white noise accounting for extra-demographic stochasticity.

To specify the measurement models, we consider three cases: Model “SEIR-VY” uses both viral load and case counts to fit the parameters, whereas models “SEIR-Y” and “SEIR-V” consider only case counts and viral load, respectively. Equations 17-22 define the likelihood of the three models along with their parameters.

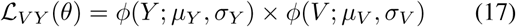

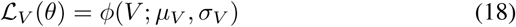

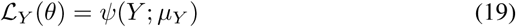

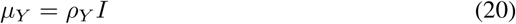

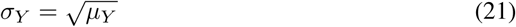

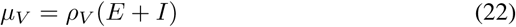

Where *ϕ*(.; *µ, σ*) is the likelihood of the normal distribution with mean *µ* and standard deviation *σ*. Similarly, *ψ*(*Y* ; *µ*) is the likelihood for the Poisson distribution.

### 2.1. Data

We use RNA data derived from our comprehensive Agent-Based Model (ABM) of the COVID-19 pandemic in Virginia. Agents in this model move randomly in a grid, and those in the same space can infect one another. Each agent is stochastically assigned a prognosis when infected, based on literature data of outcome and phase residency probabilities [26], [27]. Agents are then assigned an RNA shedding rate for each step of their disease course. Symptomatic agents start shedding RNA five days before symptom onset and peak two days before symptom onset as per literature indicated trends. Asymptomatic agents start shedding RNA when they become infectious and peak two days later. The primary input of this model is the movement probability of agents for a given step. This value is only changed for dates where significant modification to social distancing measures in Virginia occurs. The ABM fits VHD data of COVID-19 cases by date of symptom onset and is also checked against VDH data of cases by date of reporting, concurrent hospitalizations, and death rates. This ABM assumes that Virginia is a closed system, that the interactions patterns of people in Virginia can be simulated as random movement on a uniform grid, and that RNA shedding after symptoms abate is negligible.

Finally, for the research purposes of this study, we scaled down the data to the size of the city of Charlotteville, VA, and approximated the population of the city to be 50,000. We also focused on the 100 days of the early stages of the pandemic starting from March 1st, 2020 for the scope of this study.

Furthermore, we advanced the case count data to account for the reporting delays and the mismatch between the viral load data and case count data. Since most of the mean reporting delays in the literature are around five days [8], [28] we advanced the case count data by five days.

## 3. Results

In this section, we present the results of the three models, SEIR-VY, SEIR-V, and SEIR-Y. A summary of all the parameters is presented in Table 1. We assumed the initial value parameters (*η, S*_0_, *E*_0_, *I*_0_, *R*_0_) following [1]. Parameters *µ*_*SE*_, *µ*_*IR*_, and *ρ*_*Y*_ were taken from the literature. The remaining seven parameters including *β*_0_, *ι, α, σ*_*SE*_, *br, ρ*_*V*_, *σ*_*V*_ were estimated. We proceed with explaining the estimated parameters and then comparing the simulation results of the models and their forecasting quality with the data.

**Table 1:** summary parameters of the models

| Parameter | notation | value | reference |
| --- | --- | --- | --- |
| basic transmission rate | $\beta_0$ | estimated | - |
| imported infections | $\iota$ | estimated | - |
| mixing parameter | $\alpha$ | estimated | - |
| extra-demographic stochasticity shape parameter | $\sigma_{SE}$ | estimated | - |
| birth rate into susceptible | $br$ | estimated | - |
| mean shedding rate of individual (per 6 hr) | $\rho_V$ | estimated | - |
| standard deviation of shedding rate | $\sigma_V$ | estimated | - |
| reporting rate of infected agents | $\rho_Y$ | 0.14 | [9] |
| mean latent period | $1/\mu_{EI}$ | 6 days | [22] |
| mean infectious period | $1/\mu_{IR}$ | 7.7 days | [23] |
| initial fraction of the population in SEIR | $\eta$ | 0.05 | assumed |
| initial fraction of the agents in S | $S_0$ | 0.95 | assumed |
| initial fraction of the agents in E | $E_0$ | 0.04 | assumed |
| initial fraction of the agents in I | $I_0$ | 0.01 | assumed |
| initial fraction of the agents in R | $R_0$ | 0.00 | assumed |
| Total population | $P$ | 50000 | assumed |

To better evaluate the predictive power of the models, we divided the study duration of 100 days into 70 days of calibration and 30 days of projection. Models were fitted on the calibration period to find the Maximum Likelihood Estimation (MLE) parameters, 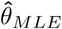. The parameter estimation was performed in R using the package “pomp” [29]. The data, along with the R codes for this study, can be found at (https://github.com/Shakeri-Lab/COVID-SEIR). Also, the MLE parameters of the three models are summarized in Table 2.

**Table 2:** summary of the MLE parameters of the models

| parameter | SEIR-VY | SEIR-V | SEIR-Y |
| --- | --- | --- | --- |
| $\beta_0$ | 0.19 | 0.14 | 0.25 |
| $\iota$ | 4.27 | 0.21 | 65.1 |
| $\alpha$ | 1.07 | 1.14 | 0.51 |
| $\sigma_{SE}$ | 1.18 | 1.11 | 5.46 |
| $br$ | 7.68 | 7.30 | 8.84 |
| $\rho_V$ | 158.9 | 150* | 150* |
| $\sigma_V$ | 3760 | 3500* | 3500* |

\*Fixed from the SEIR-VY model.

Models SEIR-VY and SEIR-V seem to be in good agreement on the optimal parameters. The only parameter that they estimate differently is the imported infections *ι*. SEIR-V estimates a trivial amount of imported infections at each timestep, while SEIR-VY estimates a non-negligible amount of imported infections, *ι* ≃ 4.27. In spite of the other two models, SEIR-Y estimates *ι* ≃ 65, which seems unreasonably high for the scale of the population under study.

Both SEIR-VY and SEIR-V estimate *α* ≃ 1 meaning a homogeneous mixing while SEIR-Y estimate *α* ≃ 0.5. *α <* 1 corresponds to some lower-level clustering in the susceptible and infectious pool resulting in a non-homogeneous transmission [30].

Shedding rates for the individuals are estimated to be ≃ 150 copies per infected case. We used the SEIR-VY to estimate the shedding rate and its standard deviation and then fixed that for the other two models. We averaged over the top 10 parameter sets with the highest log-likelihood to fix the *ρ*_*V*_ and *σ*_*V*_ for the SEIR-V and SEIR-Y.

### 3.1. Basic Reproduction Number ℛ _0_

One parameter that has a significant effect on the outcome of the model is the transmission rate. The transmission rate, *β*, plays a vital role in foreseeing the future of the epidemic and the proportion of the population that will be eventually infected. It is also linked to the basic reproduction number, which is the most important epidemiological indicator. Basic reproduction number, *ℛ*_0_, is defined as “the expected number of secondary cases produced, in a completely susceptible population, by a typical infective individual” [31]. For a model with a single infected compartment, it is equal to the product of transmission rate and the mean duration of infection [32]:

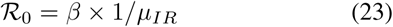

Given a fixed *µ*_*IR*_ = 0.13 we can compute the estimated basic reproduction number of the three models:

- SEIR-VY model: ℛ _0_ = 1.46.
- SEIR-V model: ℛ _0_ = 1.08.
- SEIR-Y model: ℛ _0_ = 1.92.

All of the estimated basic reproduction numbers are in agreement with previous works. For example, Turk et al. [33] estimated a basic reproduction number in the range of 1.34 to 1.79 for the city of Charlotte, NC. In another study by Kain et al. [34] reproduction numbers were calculated for some of the largest cities in the United States. Their results suggest a reproduction number of 2-4 at the beginning of the outbreak and a drastic reduction to near unity after the social distancing restrictions. Therefore, we expect a lower *ℛ*_0_ for a small city like Charlottesville, VA, which falls in the range of the estimated *ℛ*_0_ of our models.

### 3.2. Simulation Results

In order to compare the capability of the fitted models to explain viral load data and, more importantly, case count data, we first draw 1000 realizations from the three models using the estimated parameters. We then computed their mean and standard errors and plotted them against the data. Fig. 2a shows the simulation results of the models for case count for the 70 days of calibration. SEIR-VY and SEIR-V closely follow the pattern of the data, while SEIR-Y fails to match with the data. Fig. 2b also show the simulations of the models versus data. Similar to the case count, SEIR-VY and SEIR-V models can capture the trend of the viral load, while the SEIR-Y model is not detecting the peak in viral load.

**Figure 2:**
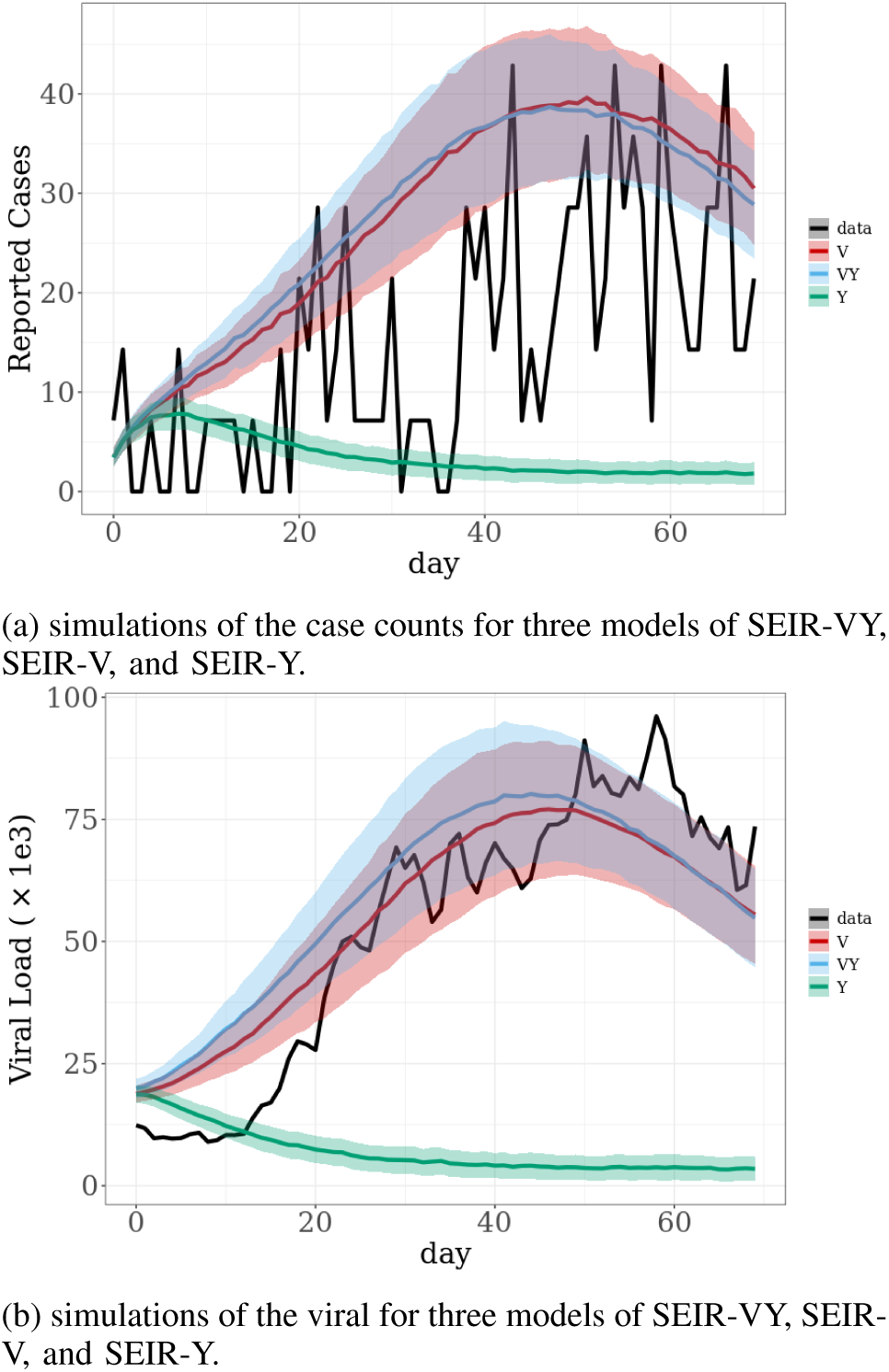
Comparison of the simulations for the first 70 days

### 3.3. Forecast Results

We also compared the prediction of three models with the data for the projection period. As a baseline, we also fitted an Autoregressive Integrated Moving Average (ARIMA) model for the case counts and compared it with the models. Using the Partial Auto Correlation Function (PACF) plots and AIC measures, we found the parameters of the ARIMA to be *p* = *q* = *d* = 1. The predictions of all four models are compared with the case count data in Fig. 3. Interestingly, SEIR-V is the best model in projecting future cases. Even the SEIR-VY model suffers from a large uncertainty, evident in its wide 95% confidence interval.

**Figure 3:**
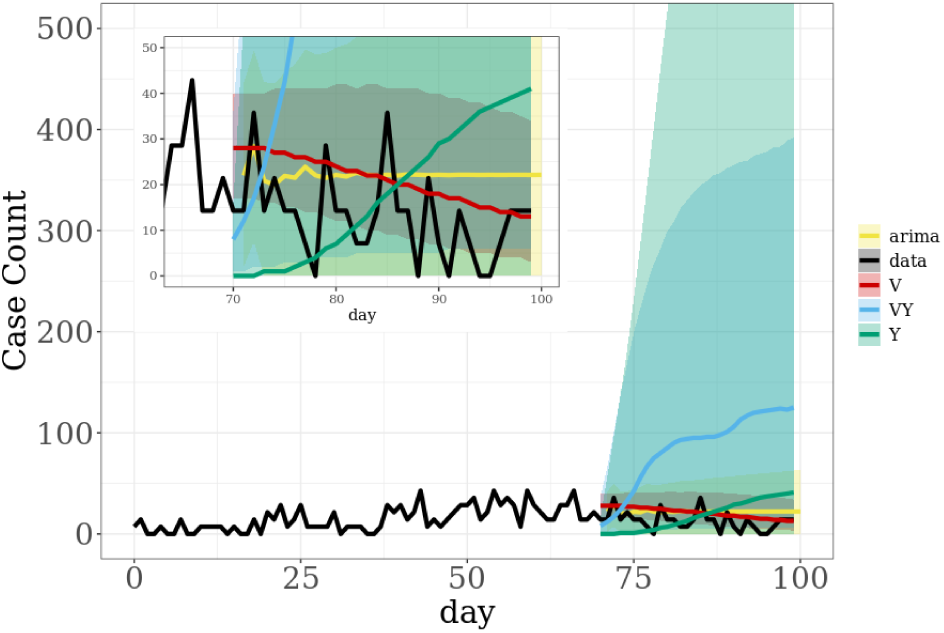
forecasts of the case counts for the four models of SEIR-VY, SEIR-V, SEIR-Y, and ARIMA. The lines are the means and the ribbons show the 95% confidence interval.

## 4. Discussion

The simulation and prediction results of the models confirm the potential of the viral load as an early indicator of an outbreak. However, the case count seems to have a lot of uncertainty surrounding it. One crucial factor is that the reporting delays vary from person to person because the latent period of the disease has a pretty wide distribution [35]. This is amplified when the number of infections is small as their mean latent period could differ considerably from the reported values by literature. Another factor that can adversely influence the reliability of the case count as an indicator is the reporting rate. Similar to the reporting delays, the reporting rate also suffers from large uncertainty that becomes even larger with a small number of infections.

On the other hand, viral load does not suffer from some of the same sources of uncertainty as the case counts. As we discussed earlier, COVID-19 is shown to have a long incubation period extending to more than ten days in some cases. However, COVID-19 patients begin excreting SARS-CoV-2 RNA in the form of urine and stool shortly after being infected and while not infectious yet [7]. Moreover, the low reporting rate of COVID-19 associated with the fact that most infections are asymptomatic or mildly symptomatic has no impact on the viral load. This is simply because all infected individuals still shed the virus, regardless of the severity of their infection.

In Fig. 3 we observe the effect of the uncertainty of the case count on the prediction of the future. Interestingly the SEIR-V model has better predictive power and much less uncertainty compared with the SEIR-VY. Out of the three SEIR models and ARIMA model, SEIR-V (red) is the only model that could predict a drop in the number of cases at the tail of the outbreak (zoomed-in panel).

## 5. Conclusion and Future Work

In this study, we implemented a SEIR model for three cases of using merely case count data, viral load data, and both. We allowed for stochasticity in the transmission rates and extra-demographic stochasticity accounting for the unforeseen events. We fitted our models using a simulation-based inference called Iterated Filtering [21]. We compared the models from three different aspects, parameter estimation, simulation, and prediction. Our results suggest that the viral load data is an informative data source for monitoring the spread of COVID-19 cases on a community level. The viral load has enough information, which enables it to approximate the number of infected cases when employed with a proper epidemiological model. Additionally, viral load data is more consistent and less uncertain, making it a critical part of COVID-19 surveillance.

Throughout this study, we made some simplifying assumptions that are important to note. The first assumption is implicit in our SEIR structure. It is a simple compartmental model that does not distinguish the infections based on the severity of their illness and whether or not they will be hospitalized. Also, we neglected the agent removals from the compartments, which could not hold in real-world scenarios and especially for larger populations. Some COVID-19 patients are shown to shed virus for a long time after the symptoms are gone [36]. Although the shedding rate decreases exponentially with time, it still can have a masking effect on newly infected cases.

Moreover, we did not take the RNA degradation through the sewer system into account. We also did not consider the social, economic, and environmental factors in our models. Factors like restriction orders can profoundly impact the behavior of the population and consequently how the infection spreads out in the community. Another notable simplification is assuming a constant transmission rate.

Many questions remain to be addressed regarding wastewater surveillance. We presented a proof of concept to show that viral load can be intelligently used to extract useful information about the outbreak. Viral load data contains information on PAMS patients and signals an outbreak days before lagging indicators like confirmed positive cases, deaths, and hospitalizations.

Future works can attempt to make the compartmental model more complex to account for different stages and variations of the infection. Nevertheless, the complexity comes at the cost of losing some interpretability, generalization possibilities and imposing more computational expenses. The impact of the different viral strains can also be studied. Other sources of data can be supplied to the model as covariates. Also, the shedding profile of the patients can be used instead of a constant shedding rate which could further improve the reliability of the inference.

## Data Availability

All of the data along with the codes in R can be found in the GitHub repository of the project.

https://github.com/Shakeri-Lab/COVID-SEIR

## Acknowledgments

The authors would like to thank the Ivy Foundation COVID-19 translational research fund for supporting this work.

## References

[1] E. H. Kaplan, D. Wang, M. Wang, A. A. Malik, A. Zulli, and J. Peccia, “Aligning SARS-CoV-2 indicators via an epidemic model: application to hospital admissions and RNA detection in sewage sludge,” Health Care Manag Sci, Oct. 2020. [Online]. Available: https://doi.org/10.1007/s10729-020-09525-1

[2] Covid-19 data dashboard. [Online]. Available: https://www.vdh.virginia.gov/coronavirus/covid-19-data-dashboard-update-schedule/

[3] E. Dong, H. Du, and L. Gardner, “An interactive web-based dashboard to track covid-19 in real time,” The Lancet infectious diseases, vol. 20, no. 5, pp. 533–534, 2020.

[4] J. Peccia, A. Zulli, D. E. Brackney, N. D. Grubaugh, E. H. Kaplan, A. Casanovas-Massana, A. I. Ko, A. A. Malik, D. Wang, M. Wang et al., “Sars-cov-2 rna concentrations in primary municipal sewage sludge as a leading indicator of covid-19 outbreak dynamics,” MedRxiv, 2020.

[5] A. Hata, R. Honda, H. Hara-Yamamura, and Y. Meuchi, “Detection of sars-cov-2 in wastewater in japan by multiple molecular assays-implication for wastewater-based epidemiology (wbe),” MedRxiv, 2020.

[6] S. Agrawal, L. Orschler, and S. Lackner, “Long-term monitoring of sars-cov-2 rna in wastewater of the frankfurt metropolitan area in southern germany,” Scientific reports, vol. 11, no. 1, pp. 1–7, 2021.

[7] T. C. Jones, G. Biele, B. Mühlemann, T. Veith, J. Schneider, J. Beheim-Schwarzbach, T. Bleicker, J. Tesch, M. L. Schmidt, L. E. Sander, F. Kurth, P. Menzel, R. Schwarzer, M. Zuchowski, J. Hofmann, A. Krumbholz, A. Stein, A. Edelmann, V. M. Corman, and C. Drosten, “Estimating infectiousness throughout SARS-CoV-2 infection course,” Science, vol. 373, no. 6551, p. eabi5273, Jul. 2021, publisher: American Association for the Advancement of Science. [Online]. Available: https://www.science.org/doi/full/10.1126/science.abi5273

[8] S. Shah, S. X. W. Gwee, J. Q. X. Ng, N. Lau, J. Koh, and J. Pang, “Wastewater surveillance to infer COVID-19 transmission: A systematic review,” Science of The Total Environment, vol. 804, p. 150060, Jan. 2022. [Online]. Available: https://www.sciencedirect.com/science/article/pii/S0048969721051354

[9] R. Li, S. Pei, B. Chen, Y. Song, T. Zhang, W. Yang, and J. Shaman, “Substantial undocumented infection facilitates the rapid dissemination of novel coronavirus (sars-cov-2),” Science, vol. 368, no. 6490, pp. 489–493, 2020.

[10] Y. Zhu, W. Oishi, C. Maruo, M. Saito, R. Chen, M. Kitajima, and D. Sano, “Early warning of COVID-19 via wastewater-based epidemiology: potential and bottlenecks,” Science of The Total Environment, vol. 767, p. 145124, May 2021. [Online]. Available: https://www.sciencedirect.com/science/article/pii/S004896972100190X

[11] S. Wurtzer, V. Marechal, J. M. Mouchel, Y. Maday, R. Teyssou, E. Richard, J. L. Almayrac, and L. Moulin, “Evaluation of lockdown impact on SARS-CoV-2 dynamics through viral genome quantification in Paris wastewaters,” Tech. Rep., May 2020, company: Cold Spring Harbor Laboratory Press Distributor: Cold Spring Harbor Laboratory Press Label: Cold Spring Harbor Laboratory Press Type: article. [Online]. Available: https://www.medrxiv.org/content/10.1101/2020.04.12.20062679v2

[12] W. Randazzo, E. Cuevas-Ferrando, R. Sanjuán, P. Domingo-Calap, and G. Sánchez, “Metropolitan wastewater analysis for COVID-19 epidemiological surveillance,” International Journal of Hygiene and Environmental Health, vol. 230, p. 113621, Sep. 2020. [Online]. Available: https://www.sciencedirect.com/science/article/pii/S1438463920305678

[13] Y. Cao and R. Francis, “On forecasting the community-level COVID-19 cases from the concentration of SARS-CoV-2 in wastewater,” Science of The Total Environment, vol. 786, p. 147451, Sep. 2021. [Online]. Available: https://www.sciencedirect.com/science/article/pii/S0048969721025225

[14] G. Medema, F. Been, L. Heijnen, and S. Petterson, “Implementation of environmental surveillance for SARS-CoV-2 virus to support public health decisions: Opportunities and challenges,” Curr Opin Environ Sci Health, vol. 17, pp. 49–71, Oct. 2020. [Online]. Available: https://www.ncbi.nlm.nih.gov/pmc/articles/PMC7528975/

[15] A. Xiao, F. Wu, M. Bushman, J. Zhang, M. Imakaev, P. R. Chai, C. Duvallet, N. Endo, T. B. Erickson, F. Armas, B. Arnold, H. Chen, F. Chandra, N. Ghaeli, X. Gu, W. P. Hanage, W. L. Lee, M. Matus, K. A. McElroy, K. Moniz, S. F. Rhode, J. Thompson, and E. J. Alm, “Metrics to relate COVID-19 wastewater data to clinical testing dynamics,” Infectious Diseases (except HIV/AIDS), preprint, Jun. 2021. [Online]. Available: http://medrxiv.org/lookup/doi/10.1101/2021.06.10.21258580

[16] C. S. McMahan, S. Self, L. Rennert, C. Kalbaugh, D. Kriebel, D. Graves, J. A. Deaver, S. Popat, T. Karanfil, and D. L. Freedman, “COVID-19 Wastewater Epidemiology: A Model to Estimate Infected Populations,” Tech. Rep., Nov. 2020, company: Cold Spring Harbor Laboratory Press Distributor: Cold Spring Harbor Laboratory Press Label: Cold Spring Harbor Laboratory Press Type: article. [Online]. Available: https://www.medrxiv.org/content/10.1101/2020.11.05.20226738v1

[17] S. Nourbakhsh, A. Fazil, M. Li, C. S. Mangat, S. W. Peterson, J. Daigle, S. Langner, J. Shurgold, P. D’Aoust, R. Delatolla, E. Mercier, X. Pang, B. E. Lee, R. Stuart, S. Wijayasri, and D. Champredon, “A Wastewater-Based Epidemic Model for SARS-CoV-2 with Application to Three Canadian Cities,” Epidemiology, preprint, Jul. 2021. [Online]. Available: http://medrxiv.org/lookup/doi/10.1101/2021.07.19.21260773

[18] C. Bretó, D. He, E. L. Ionides, and A. A. King, “Time series analysis via mechanistic models,” Ann. Appl. Stat., vol. 3, no. 1, Mar. 2009, 0802.0021. [Online]. Available: http://arxiv.org/abs/0802.0021

[19] E. L. Ionides, D. Nguyen, Y. Atchadé, S. Stoev, and A. A. King, “Inference for dynamic and latent variable models via iterated, perturbed Bayes maps,” Proc Natl Acad Sci U S A, vol. 112, no. 3, pp. 719–724, Jan. 2015. [Online]. Available: https://www.ncbi.nlm.nih.gov/pmc/articles/PMC4311819/

[20] E. L. Ionides, A. Bhadra, Y. Atchadé, and A. King, “Iterated filtering,” The Annals of Statistics, vol. 39, no. 3, pp. 1776–1802, 2011.

[21] E. L. Ionides, C. Breto, and A. A. King, “Inference for nonlinear dynamical systems,” Proceedings of the National Academy of Sciences, vol. 103, no. 49, pp. 18 438–18 443, Dec. 2006. [Online]. Available: http://www.pnas.org/cgi/doi/10.1073/pnas.0603181103

[22] Covid-19 pandemic planning scenarios. [Online]. Available: https://www.cdc.gov/coronavirus/2019-ncov/cases-updates/burden.html

[23] A. W. Byrne, D. McEvoy, A. B. Collins, K. Hunt, M. Casey, A. Barber, F. Butler, J. Griffin, E. A. Lane, C. McAloon et al., “Inferred duration of infectious period of sars-cov-2: rapid scoping review and analysis of available evidence for asymptomatic and symptomatic covid-19 cases,” BMJ open, vol. 10, no. 8, p. e039856, 2020.

[24] Estimated covid-19 burden. [Online]. Available: https://www.cdc.gov/coronavirus/2019-ncov/hcp/planning-scenarios.html

[25] D. He, E. L. Ionides, and A. A. King, “Plug-and-play inference for disease dynamics: measles in large and small populations as a case study,” Journal of The Royal Society Interface, vol. 7, no. 43, pp. 271–283, Feb. 2010, publisher: Royal Society. [Online]. Available: https://royalsocietypublishing.org/doi/10.1098/rsif.2009.0151

[26] V. A. Avanzato, M. J. Matson, S. N. Seifert, R. Pryce, B. N. Williamson, S. L. Anzick, K. Barbian, S. D. Judson, E. R. Fischer, C. Martens et al., “Case study: prolonged infectious sars-cov-2 shedding from an asymptomatic immunocompromised individual with cancer,” Cell, vol. 183, no. 7, pp. 1901–1912, 2020.

[27] S. Lee, T. Kim, E. Lee, C. Lee, H. Kim, H. Rhee, S. Y. Park, H.-J. Son, S. Yu, J. W. Park et al., “Clinical course and molecular viral shedding among asymptomatic and symptomatic patients with sars-cov-2 infection in a community treatment center in the republic of korea,” JAMA internal medicine, vol. 180, no. 11, pp. 1447–1452, 2020.

[28] J. E. Harris, “Overcoming reporting delays is critical to timely epidemic monitoring: The case of covid-19 in new york city,” MedRxiv, 2020.

[29] A. A. King, D. Nguyen, and E. L. Ionides, “Statistical Inference for Partially Observed Markov Processes via the R Package pomp,” J. Stat. Soft., vol. 69, no. 12, 2016, 1509.00503. [Online]. Available: http://arxiv.org/abs/1509.00503

[30] O. N. Bjørnstad, B. F. Finkenstädt, and B. T. Grenfell, “Dynamics of measles epidemics: estimating scaling of transmission rates using a time series sir model,” Ecological monographs, vol. 72, no. 2, pp. 169–184, 2002.

[31] O. Diekmann, J. A. P. Heesterbeek, and J. A. Metz, “On the definition and the computation of the basic reproduction ratio r 0 in models for infectious diseases in heterogeneous populations,” Journal of mathematical biology, vol. 28, no. 4, pp. 365–382, 1990.

[32] P. van den Driessche and J. Watmough, “Reproduction numbers and sub-threshold endemic equilibria for compartmental models of disease transmission,” Mathematical Biosciences, vol. 180, no. 1, pp. 29–48, Nov. 2002. [Online]. Available: https://www.sciencedirect.com/science/article/pii/S0025556402001086

[33] P. J. Turk, S.-H. Chou, M. A. Kowalkowski, P. P. Palmer, J. S. Priem, M. D. Spencer, Y. J. Taylor, and A. D. McWilliams, “Modeling covid-19 latent prevalence to assess a public health intervention at a state and regional scale: Retrospective cohort study,” JMIR public health and surveillance, vol. 6, no. 2, p. e19353, 2020.

[34] M. P. Kain, M. L. Childs, A. D. Becker, and E. A. Mordecai, “Chopping the tail: How preventing superspreading can help to maintain COVID-19 control,” Epidemics, vol. 34, p. 100430, Mar. 2021. [Online]. Available: https://www.sciencedirect.com/science/article/pii/S1755436520300487

[35] H. Xin, Y. Li, P. Wu, Z. Li, E. H. Lau, Y. Qin, L. Wang, B. J. Cowling, T. Tsang, and Z. Li, “Estimating the latent period of coronavirus disease 2019 (covid-19),” Clinical Infectious Diseases: an Official Publication of the Infectious Diseases Society of America, 2021.

[36] Y. Wu, C. Guo, L. Tang, Z. Hong, J. Zhou, X. Dong, H. Yin, Q. Xiao, Y. Tang, X. Qu, L. Kuang, X. Fang, N. Mishra, J. Lu, H. Shan, G. Jiang, and X. Huang, “Prolonged presence of SARS-CoV-2 viral RNA in faecal samples,” The Lancet Gastroenterology & Hepatology, vol. 5, no. 5, pp. 434–435, May 2020, publisher: Elsevier. [Online]. Available: https://www.thelancet.com/journals/langas/article/PIIS2468-1253(20)30083-2/abstract

